# Global emergence of resistance to fluconazole and voriconazole in *Candida parapsilosis* in tertiary hospitals in Spain during the COVID-19 pandemic

**DOI:** 10.1101/2022.06.06.22275514

**Authors:** Oscar Zaragoza, Laura Alcázar-Fuoli, Nuria Trevijano-Contador, Alba Torres-Cano, Cristina Carballo-González, Mireia Puig-Asensio, María Teresa Martín-Gómez, Emilio Jiménez-Martínez, Daniel Romero, Francesc Xavier Nuvials, Roberto Olmos-Arenas, María Clara Moretó-Castellsagué, Lucía Fernández-Delgado, Graciela Rodríguez-Sevilla, María-Mercedes Aguilar-Sánchez, Josefina Ayats-Ardite, Carmen Ardanuy-Tisaire, Isabel Sanchez-Romero, María Muñoz-Algarra, Paloma Merino-Amador, Fernando González-Romo, Gregoria Megías-Lobón, Jose Angel García-Campos, María Ángeles Mantecón-Vallejo, Maria Teresa Durán-Valle, Arturo Manuel Fraile-Torres, María Pía Roiz-Mesones, Isabel Lara-Plaza, Ana Perez de Ayala, María Simón-Sacristán, Ana Collazos-Blanco, Teresa Nebreda-Mayoral, Gabriel March-Roselló

## Abstract

**Background:** *Candida parapsilosis* is a frequent cause of candidemia worldwide. Its incidence is associated with the use of medical implants, such as central venous catheters or parenteral nutrition. This species has reduced susceptibility to echinocandins and is susceptible to polyenes and azoles. Multiple outbreaks caused by fluconazole non-susceptible strains have been reported recently. A similar trend has been observed among the *C. parapsilosis* isolates received in the last two years at the Spanish Mycology Reference Laboratory.

**Methods:** Yeast were identified by molecular biology and antifungal susceptibility testing was performed using EUCAST protocol. *ERG11* gene was sequenced to identify resistance mechanisms, and typification was carried out by microsatellite analysis.

**Results:** We examined the susceptibility profile of the *C. parapsilosis* isolates available at our Reference Laboratory since 2000 (around 1,300 strains). During the last two years, the number of isolates with acquired resistance to fluconazole and voriconazole has increased in at least eight different Spanish hospitals. Typification of the isolates revealed that some prevalent clones had spread through several hospitals of the same geographical region. One of these clones was found in hospitals from the region of Catalonia, another in hospitals from Madrid and Burgos, and two other different genotypes from Santander.

**Conclusions:** Our data suggests that the epidemiological situation caused by the COVID-19 pandemic might have induced a selection of fluconazole-resistant *C. parapsilosis* isolates that were already present at the hospitals. Further measures must be taken to avoid the establishment of clinical outbreaks that could threaten the life of infected patients.

## INTRODUCTION

*Candida parapsilosis* is an opportunistic pathogenic yeast able to cause invasive diseases such as candidemia. Worldwide, it is the third cause of bloodborne yeast infection after *C. albicans* and *C. glabrata*, although in some countries, its incidence is higher and above *C. glabrata* [1-4]. Neonates, as well as indwelling parenteral nutrition and central nervous catheters have been associated to a higher risk of infection [5, 6]. Besides sporadic infections, *C. parapsilosis* is well known to cause nosocomial outbreaks through direct and indirect contact via the hands of health care workers and through contaminated patient care equipment.

*Candida parapsilosis* exhibits a reduced natural in vitro susceptibility to echinocandins [7], so the main therapeutic options for invasive infections due to this species are the triazoles, mainy fluconazole or, alternatively, polyenes. Acquired resistance to fluconazole in *C. parapsilosis* is a rare phenomenon, being less than 5% of isolates in different epidemiological studies [2, 7-10]. In recent years, however, a steady increase of resistance has been observed worldwide, mostly in the context of nosocomial outbreaks [11-20]. In many cases, these outbreaks are monoclonal, and are associated to mutations in *ERG11* (mainly with the Y132F mutation), overexpression of efflux pumps (as Mdr1 and Cdr1) and mutations in *MRR1*, which encodes a transcription factor that regulates the expression of some efflux pumps [11-13, 16, 18, 21, 22].

The National Centre for Microbiology from Instituto de Salud Carlos III (CNM-ISCIII, Madrid, Spain) acts as a national reference center for clinically isolated fungi, providing services such as genotyping and confirmation of antifungal susceptibility profiles by the EUCAST standardized methodology. Since 2020 a significant increase in the number of fluconazole non-susceptible (FNS) *C. parapsilosis* isolates received was noted, most of them coming from tertiary hospitals across the country reporting to have a strong epidemiological suspicion of ongoing outbreaks.

The aim of this work was to describe the antifungal susceptibility profile of all the *C. parapsilosis* isolates received in the Spanish Mycology Reference Laboratory (SMRL) since 2000 to get insights about susceptibility profile and appearance of resistance in this species. Typing analysis confirmed genetic relatedness between isolates and suggested that in Spain there could be an expansion of *C. parapsilosis* resistant isolates among tertiary care hospitals. The fact that this expansion overlaps with the impact of the COVID-19 pandemic, highlights a worrisome situation in which resistance to azoles in *C. parapsilosis* could be emerging worldwide.

## MATERIAL AND METHODS

### Media and strains identification

The isolates were primarily isolated, identified and screened for fluconazole non-susceptibility at the local laboratories following the routine methodologies of each center. Isolates sent to the CNM-ISCIII since 2000 and identified as *C. parapsilosis* were subcultivated onto Sabouraud solid or liquid medium. Identification was confirmed by sequencing the internal transcribed spacer 1 (ITS1) and ITS2 regions from the ribosomal DNA as previously in [23].

### Antifungal susceptibility

Antifungal susceptibility testing was performed following EUCAST protocol (RPMI 1640 medium (Merck, Sigma-Aldrich) buffered with MOPS (Merck, Sigma-Aldrich) at pH 7 and supplemented with 2% glucose (Merck, Sigma-Aldrich), [24]).The following antifungals were tested in the concentration range indicated in brackets: Amphotericin B (AmB, Merck, Sigma-Aldrich, 16-0.03 mg/L), flucytosine (64-0.125 mg/L), fluconazole (FLC, Merck, Sigma-Aldrich, 64-0.125 mg/L), itraconazole (ITZ, Janssen Pharmaceutical Research and Development, 8-0.016 mg/L), voriconazole (VOR, Pfizer Pharmaceutical Group, 8-0.016 mg/L), posaconazole (POS, Merck, Sigma-Aldrich, 8-0.016 mg/L), isavuconazole (ISV, Pfizer Pharmaceutical Group, 8-0.016 mg/L), caspofungin (CSP, Merck, Sigma-Aldrich, 16-0.016 mg/L), micafungin (MICA, Astellas Pharma Inc, 2-0.004 mg/L) and anidulafungin (ANID, Pfizer Pharmaceutical Group, 4-0.008 mg/L). The Minimal inhibitory concentration (MIC) was defined as the concentration that caused 50% of growth inhibition compared to the control well without antifungal, except for amphotericin B (90%). Strains were categorized as susceptible (S), resistant (R) or intermediate (I, susceptible, increased exposure) following the breakpoints established by EUCAST (see https://www.eucast.org/astoffungi/clinicalbreakpointsforantifungals, document from February 4th, 2020). Control strains *C. parapsilosis* ATCC 22019 and *C. krusei* ATCC 6258 were included in all the assays.

### Sequencing of the *ERG11* gene

To identify mutations at the *ERG11* gene, different primers were designed (see table 1). The whole gene was amplified using oligonucleotides 01_F and 02_R using the following PCR conditions: 94°C for 2 min and 35 cycles of amplification (94°C for 30 s, 56°C for 45 s and 72°C for 2 min) followed by a 1 final cycle of 5 min of 72°C.

**Table 1.**
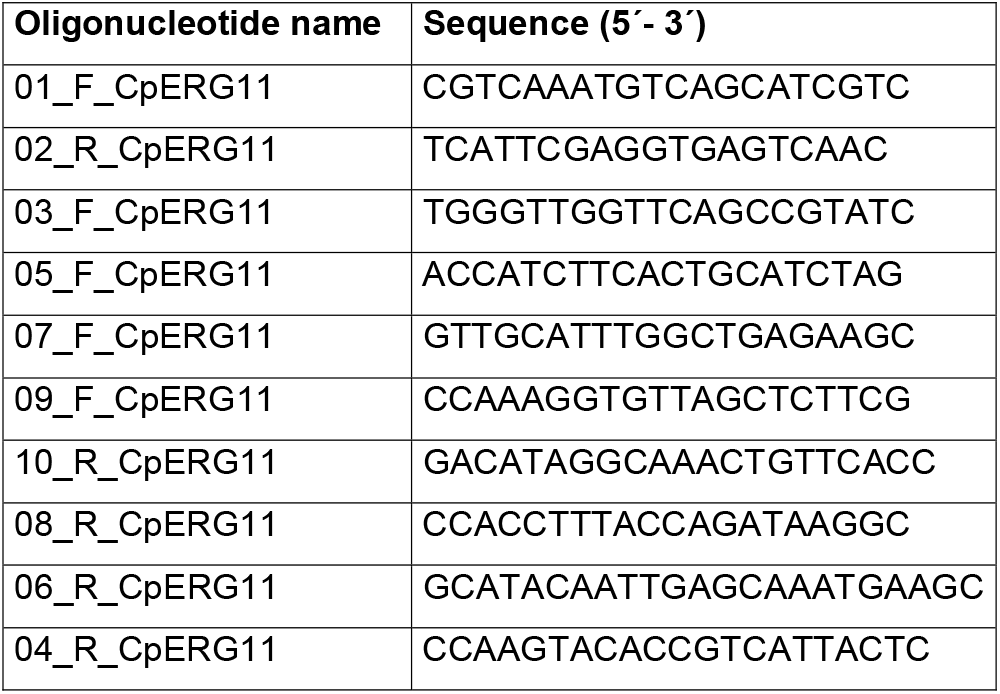
Oligonucleotides designed to sequence the *Candida parapsilosis ERG11* gene (F, forward; R, reverse).

The PCR products were purified with ExoStart kit. Sanger sequencing was performed using all the oligonucleotides described in table 1, and analyzed with Seqman software (DNA Lasergene 12 package).

### Microsatellite typing

A panel of four short tandem repeat (STR) markers was used for genotyping the *C. parapsilosis* isolates. Three trinucleotide repeat and one hexanucleotide repeat markers described by Diab-Elschahawi [25] were independently amplified by PCR. Amplifications reactions were performed in a final volume of 20 µl for markers 3A, 3B and 6A, containing 1 ng of genomic DNA, 0.5 µM amplification primers, 120 µM of dNTPS, 1.25 mM MgCl_2_ and 1 U of Amplitaq™ DNA (Applied Biosystems). The PCR conditions used were: initial denaturalization for 5 min at 95 °C, followed by 30 cycles of 15 s of denaturalization at 95 °C, 1 min of annealing at 62 °C, and 1 min of extension at 72 °C. A final incubation of 7 min at 72 °C was included in the protocol. The PCR conditions were optimized for the “3C” marker. In this case, amplifications reactions were performed in a final volume of 50 µl, containing a 1 ng of DNA, 0.2 µM amplification primers, 0.05 mM of dNTPS, 0.3 mM MgCl_2_ and 1 U of Amplitaq™ DNA (Applied Biosystems). PCR conditions for 3C marker were as follows: initial denaturalization for 5 min at 94 °C, followed by 35 cycles of 30 s of denaturalization at 94 °C, 45 s of annealing at 60 °C, and 1 min of extension at 72 °C followed of 5 min at 72 °C. Then, 10 µl of the amplification products were put on PCR Plate 96 semi-skirted (Eppendorf) and purified with AMpure XP (Beckman Coulter) using SPRI beads technology in an Eppendorf ep Motion 5075 (Eppendorf). Finally, a 1 µl aliquot of PCR product was added to a 9 µl of Formamide and to a 1 µl of internal size marker GeneScan™ 500 ROX™ (Applied Biosystems). After denaturalization of the samples at 95 °C for 3 min and rapid cooling to 4 °C, they were run onto a AB3730XL DNA analyzer (Applied Biosystems). Allele sizes analysis was performed with the Peak scanner software (Applied Biosystems) and according the internal lane size standard GeneScan™ 500 ROX™.

Similarities between genotypes were visualized by constructing a minimum spanning tree using InfoQuest FP, version 4.5 (Applied Maths, St.-Martens-Latem, Belgium), treating the data as categorical information.

### Data analysis and Statistics

MIC analysis was performed with SPSS software. For each year, the distribution of MICs was reported. We also calculated the geometric mean of the MICs values, the median, and the minimal and maximal values of the distributions.

## RESULTS AND DISCUSSION

We collected all the isolates available at our laboratory (SMRL), and analysed the evolution of the antifungal susceptibility pattern from 2000 to 2021. A total of 1,301 isolates were studied. As shown in table 1, resistance to fluconazole remained low (3-7%) among the isolates from our collection until 2016. However, a dramatic change in this resistance rate among the isolates received at the Reference Laboratory was noted thereafter, being particularly notable from 2019 onwards. Throughout the latter period, the percentage of fluconazole resistance significantly increased (27% in 2019, around 60% in 2020 and 2021, see table 2) as compared to previous years. This trend was also observed for voriconazole (Table 3). Before 2019, the voriconazole resistance rate was below 2%, but since 2020, the percentage of susceptible increased exposure (I, MIC = 0.25 mg/L) and resistant strains (MIC>0.25 mg/L) increased up to around 60% among the strains received at the laboratory.

**Table 2:** Distribution of the percentage of MICs to FLUCONAZOLE of *C. parapsilosis* strains received at the SMRL since 2000. The table include the number of strains analysed each year, and the % of susceptible (S), susceptible increased exposure (I) and resistant (R) isolates.

|  | MIC (mg/L) |  |  |  |  |  |  |  |  |  |  |  |  |  |  |
| --- | --- | --- | --- | --- | --- | --- | --- | --- | --- | --- | --- | --- | --- | --- | --- |
|  | SUSCEPTIBLE |  |  |  |  | I | RESISTANT |  |  |  |  |  |  |  |  |
| Year | 0.125 | 0.25 | 0.5 | 1 | 2 | 4 | 8 | 16 | 32 | 64 | >64 | N | % S | % I | % R |
| 2000 | 2 | 33 | 35 | 16 | 12 | 2 | 0 | 0 | 0 | 0 | 0 | 43 | 98 | 2 | 0 |
| 2001 | 1 | 30 | 53 | 12 | 1 | 3 | 0 | 0 | 0 | 0 | 0 | 74 | 97 | 3 | 0 |
| 2002 | 8 | 36 | 48 | 5 | 3 | 0 | 0 | 0 | 0 | 1 | 0 | 80 | 99 | 0 | 1 |
| 2003 | 11 | 40 | 41 | 5 | 1 | 0 | 1 | 1 | 0 | 0 | 0 | 115 | 98 | 0 | 2 |
| 2004 | 7 | 35 | 44 | 9 | 5 | 0 | 0 | 0 | 0 | 0 | 0 | 75 | 100 | 0 | 0 |
| 2005 | 18 | 41 | 32 | 6 | 1 | 0 | 0 | 0 | 1 | 0 | 0 | 82 | 99 | 0 | 1 |
| 2006 | 3 | 27 | 49 | 17 | 3 | 1 | 0 | 0 | 0 | 0 | 0 | 71 | 99 | 1 | 0 |
| 2007 | 3 | 24 | 56 | 8 | 6 | 0 | 1 | 0 | 0 | 0 | 1 | 71 | 97 | 0 | 3 |
| 2008 | 3 | 27 | 55 | 5 | 3 | 1 | 1 | 0 | 2 | 0 | 1 | 92 | 95 | 1 | 4 |
| 2009 | 0 | 12 | 78 | 4 | 0 | 0 | 4 | 1 | 0 | 0 | 0 | 73 | 95 | 0 | 5 |
| 2010 | 9 | 11 | 64 | 9 | 2 | 0 | 2 | 4 | 0 | 0 | 0 | 55 | 95 | 0 | 5 |
| 2011 | 5 | 14 | 59 | 14 | 0 | 0 | 9 | 0 | 0 | 0 | 0 | 22 | 91 | 0 | 9 |
| 2012 | 14 | 14 | 50 | 23 | 0 | 0 | 0 | 0 | 0 | 0 | 0 | 22 | 100 | 0 | 0 |
| 2013 | 0 | 14 | 57 | 21 | 0 | 0 | 7 | 0 | 0 | 0 | 0 | 14 | 93 | 0 | 7 |
| 2014 | 15 | 12 | 46 | 23 | 0 | 0 | 0 | 0 | 0 | 4 | 0 | 26 | 96 | 0 | 4 |
| 2015 | 0 | 2 | 61 | 37 | 0 | 0 | 0 | 0 | 0 | 0 | 0 | 46 | 100 | 0 | 0 |
| 2016 | 0 | 17 | 48 | 17 | 9 | 4 | 4 | 0 | 0 | 0 | 0 | 23 | 91 | 4 | 4 |
| 2017 | 9 | 22 | 39 | 4 | 4 | 4 | 4 | 9 | 4 | 0 | 0 | 23 | 78 | 4 | 17 |
| 2018 | 0 | 15 | 85 | 0 | 0 | 0 | 0 | 0 | 0 | 0 | 0 | 13 | 100 | 0 | 0 |
| 2019 | 18 | 9 | 9 | 18 | 9 | 9 | 0 | 0 | 9 | 0 | 18 | 11 | 64 | 9 | 27 |
| 2020 | 3 | 6 | 20 | 6 | 0 | 2 | 8 | 20 | 12 | 17 | 5 | 66 | 35 | 1.5 | 63.5 |
| 2021 | 3 | 13 | 12 | 1 | 4 | 3 | 10 | 36 | 11 | 5 | 1 | 204 | 33 | 3 | 64 |

Cells in orange: Mode (most frequent value) In yellow: left and right values to the mode

**Table 3:** Distribution of the percentage of MIC to VORICONAZOLE of *C. parapsilosis* strains received at the SMRL since 2000. The table include the number of strains analysed each year, and the % of susceptible (S), susceptible increased exposure (I) and resistant (R) isolates.

| Year | MIC (mg/L) |  |  |  |  |  |  |  |  |  |  | N | % S | % I | % R |
| --- | --- | --- | --- | --- | --- | --- | --- | --- | --- | --- | --- | --- | --- | --- | --- |
|  | SUSCEPTIBLE |  |  |  | I | RESISTANT |  |  |  |  |  |  |  |  |  |
|  | 0.016 | 0.031 | 0.06 | 0.125 | 0.25 | 0.5 | 1 | 2 | 4 | 8 | >8 |  |  |  |  |
| 2000 | 79 | 14 | 7 | 0 | 0 | 0 | 0 | 0 | 0 | 0 | 0 | 42 | 100 | 0 | 0 |
| 2001 | 73 | 23 | 1 | 3 | 0 | 0 | 0 | 0 | 0 | 0 | 0 | 74 | 100 | 0 | 0 |
| 2002 | 85 | 14 | 0 | 0 | 0 | 1 | 0 | 0 | 0 | 0 | 0 | 80 | 99 | 0 | 1 |
| 2003 | 88 | 11 | 0 | 1 | 0 | 0 | 0 | 0 | 0 | 0 | 0 | 94 | 100 | 0 | 0 |
| 2004 | 96 | 4 | 0 | 0 | 0 | 0 | 0 | 0 | 0 | 0 | 0 | 75 | 100 | 0 | 0 |
| 2005 | 93 | 6 | 0 | 0 | 1 | 0 | 0 | 0 | 0 | 0 | 0 | 82 | 99 | 1 | 0 |
| 2006 | 94 | 4 | 1 | 0 | 0 | 0 | 0 | 0 | 0 | 0 | 0 | 71 | 100 | 0 | 0 |
| 2007 | 89 | 6 | 4 | 0 | 0 | 0 | 0 | 0 | 0 | 0 | 1 | 71 | 99 | 0 | 1 |
| 2008 | 90 | 4 | 0 | 2 | 0 | 1 | 1 | 1 | 0 | 0 | 0 | 92 | 97 | 0 | 3 |
| 2009 | 89 | 5 | 1 | 3 | 1 | 0 | 0 | 0 | 0 | 0 | 0 | 73 | 99 | 1 | 0 |
| 2010 | 85 | 4 | 4 | 5 | 2 | 0 | 0 | 0 | 0 | 0 | 0 | 55 | 98 | 2 | 0 |
| 2011 | 86 | 9 | 0 | 5 | 0 | 0 | 0 | 0 | 0 | 0 | 0 | 22 | 100 | 0 | 0 |
| 2012 | 77 | 18 | 5 | 0 | 0 | 0 | 0 | 0 | 0 | 0 | 0 | 22 | 100 | 0 | 0 |
| 2013 | 79 | 14 | 0 | 7 | 0 | 0 | 0 | 0 | 0 | 0 | 0 | 14 | 100 | 0 | 0 |
| 2014 | 69 | 23 | 4 | 0 | 0 | 0 | 4 | 0 | 0 | 0 | 0 | 26 | 96 | 0 | 4 |
| 2015 | 89 | 9 | 0 | 2 | 0 | 0 | 0 | 0 | 0 | 0 | 0 | 46 | 100 | 0 | 0 |
| 2016 | 70 | 13 | 13 | 4 | 0 | 0 | 0 | 0 | 0 | 0 | 0 | 23 | 100 | 0 | 0 |
| 2017 | 57 | 26 | 0 | 9 | 0 | 4 | 4 | 0 | 0 | 0 | 0 | 23 | 91 | 0 | 9 |
| 2018 | 87 | 0 | 0 | 0 | 0 | 13 | 0 | 0 | 0 | 0 | 0 | 15 | 87 | 0 | 13 |
| 2019 | 36 | 36 | 0 | 0 | 0 | 9 | 0 | 18 | 0 | 0 | 0 | 11 | 73 | 0 | 27 |
| 2020 | 27 | 8 | 0 | 2 | 2 | 24 | 36 | 2 | 0 | 0 | 0 | 66 | 36 | 2 | 62 |
| 2021 | 28 | 2 | 3 | 5 | 25 | 27 | 7 | 1 | 1 | 0 | 0 | 204 | 38 | 25 | 37 |

Cells in orange: Mode (most frequent value)
In yellow: left and right values to the mode

Regarding itraconazole and posaconazole, there was a slight trend to higher MICs, but they were still categorized as susceptible (Table 4 and 5). Only three isolates were fully resistant to fluconazole, voriconazole, itraconazole and posaconazole. For isavuconazole, although there are not breakpoints to define resistant strains, an increase in the MICs among the isolates received since 2020 was found. The isavuconazole modal MIC rose from 0.016 mg/L before 2020 to 0.06 mg/L later on (table 6), similarly to what was observed for itraconazole and posaconazole.

**Table 4:** Distribution of the percentage of MIC to ITRACONAZOLE of *C. parapsilosis* strains received at the SMRL since 2000.

|  | MIC (mg/L) |  |  |  |  |  |  |  |  |  |  |  |  |  |
| --- | --- | --- | --- | --- | --- | --- | --- | --- | --- | --- | --- | --- | --- | --- |
|  | SUSCEPTIBLE |  |  |  | RESISTANT |  |  |  |  |  |  |  |  |  |
| YEAR | 0.016 | 0.031 | 0.06 | 0.125 | 0.25 | 0.5 | 1 | 2 | 4 | 8 | >8 | N | % S | % R |
| 2000 | 58 | 30 | 9 | 2 | 0 | 0 | 0 | 0 | 0 | 0 | 0 | 43 | 100 | 0 |
| 2001 | 30 | 55 | 11 | 4 | 0 | 0 | 0 | 0 | 0 | 0 | 0 | 74 | 100 | 0 |
| 2002 | 25 | 56 | 18 | 0 | 1 | 0 | 0 | 0 | 0 | 0 | 0 | 80 | 99 | 1 |
| 2003 | 50 | 36 | 12 | 1 | 0 | 1 | 0 | 0 | 0 | 0 | 0 | 115 | 99 | 1 |
| 2004 | 61 | 31 | 7 | 1 | 0 | 0 | 0 | 0 | 0 | 0 | 0 | 75 | 100 | 0 |
| 2005 | 52 | 41 | 6 | 0 | 0 | 0 | 0 | 0 | 0 | 0 | 0 | 82 | 100 | 0 |
| 2006 | 73 | 20 | 6 | 1 | 0 | 0 | 0 | 0 | 0 | 0 | 0 | 71 | 100 | 0 |
| 2007 | 63 | 25 | 8 | 1 | 0 | 0 | 0 | 0 | 0 | 0 | 1 | 71 | 99 | 1 |
| 2008 | 52 | 40 | 4 | 0 | 2 | 1 | 0 | 0 | 0 | 0 | 0 | 92 | 97 | 3 |
| 2009 | 78 | 21 | 0 | 1 | 0 | 0 | 0 | 0 | 0 | 0 | 0 | 73 | 100 | 0 |
| 2010 | 95 | 0 | 2 | 2 | 2 | 0 | 0 | 0 | 0 | 0 | 0 | 55 | 98 | 2 |
| 2011 | 86 | 9 | 5 | 0 | 0 | 0 | 0 | 0 | 0 | 0 | 0 | 22 | 100 | 0 |
| 2012 | 95 | 5 | 0 | 0 | 0 | 0 | 0 | 0 | 0 | 0 | 0 | 22 | 100 | 0 |
| 2013 | 71 | 29 | 0 | 0 | 0 | 0 | 0 | 0 | 0 | 0 | 0 | 14 | 100 | 0 |
| 2014 | 65 | 23 | 4 | 0 | 4 | 4 | 0 | 0 | 0 | 0 | 0 | 26 | 92 | 8 |
| 2015 | 72 | 28 | 0 | 0 | 0 | 0 | 0 | 0 | 0 | 0 | 0 | 46 | 100 | 0 |
| 2016 | 35 | 22 | 30 | 13 | 0 | 0 | 0 | 0 | 0 | 0 | 0 | 23 | 100 | 0 |
| 2017 | 26 | 35 | 22 | 9 | 9 | 0 | 0 | 0 | 0 | 0 | 0 | 23 | 91 | 9 |
| 2018 | 7 | 20 | 67 | 7 | 0 | 0 | 0 | 0 | 0 | 0 | 0 | 15 | 100 | 0 |
| 2019 | 9 | 18 | 36 | 27 | 9 | 0 | 0 | 0 | 0 | 0 | 0 | 11 | 91 | 9 |
| 2020 | 15 | 20 | 41 | 23 | 2 | 0 | 0 | 0 | 0 | 0 | 0 | 66 | 98 | 2 |
| 2021 | 23 | 28 | 22 | 20 | 6 | 1 | 1 | 0 | 0 | 0 | 0 | 204 | 89 | 11 |

Cells in orange: Mode (most frequent value) In yellow: left and right values to the mode

**Table 5:** Distribution of the percentage of MIC to POSACONAZOLE of *C. parapsilosis* strains received at the SMRL since 2000

|  | MIC (mg/L) |  |  |  |  |  |  |  |  |  |  |  |  |  |
| --- | --- | --- | --- | --- | --- | --- | --- | --- | --- | --- | --- | --- | --- | --- |
|  | SUSCEPTIBLE |  |  | RESISTANT |  |  |  |  |  |  |  |  |  |  |
| YEAR | 0.016 | 0.031 | 0.06 | 0.125 | 0.25 | 0.5 | 1 | 2 | 4 | 8 | >8 | N | % S | % R |
| 2001 | 55 | 45 | 0 | 0 | 0 | 0 | 0 | 0 | 0 | 0 | 0 | 11 | 100 | 0 |
| 2002 | 65 | 31 | 4 | 0 | 0 | 0 | 0 | 0 | 0 | 0 | 0 | 80 | 100 | 0 |
| 2003 | 80 | 19 | 1 | 0 | 0 | 0 | 0 | 0 | 0 | 0 | 0 | 94 | 100 | 0 |
| 2004 | 83 | 17 | 0 | 0 | 0 | 0 | 0 | 0 | 0 | 0 | 0 | 75 | 100 | 0 |
| 2005 | 65 | 33 | 2 | 0 | 0 | 0 | 0 | 0 | 0 | 0 | 0 | 82 | 100 | 0 |
| 2006 | 87 | 13 | 0 | 0 | 0 | 0 | 0 | 0 | 0 | 0 | 0 | 70 | 100 | 0 |
| 2007 | 87 | 11 | 0 | 0 | 0 | 0 | 0 | 0 | 1 | 0 | 0 | 71 | 99 | 1 |
| 2008 | 70 | 26 | 1 | 1 | 1 | 1 | 0 | 0 | 0 | 0 | 0 | 92 | 97 | 3 |
| 2009 | 81 | 19 | 0 | 0 | 0 | 0 | 0 | 0 | 0 | 0 | 0 | 73 | 100 | 0 |
| 2010 | 74 | 20 | 2 | 2 | 2 | 0 | 0 | 0 | 0 | 0 | 0 | 54 | 96 | 4 |
| 2011 | 95 | 5 | 0 | 0 | 0 | 0 | 0 | 0 | 0 | 0 | 0 | 22 | 100 | 0 |
| 2012 | 91 | 9 | 0 | 0 | 0 | 0 | 0 | 0 | 0 | 0 | 0 | 22 | 100 | 0 |
| 2013 | 93 | 7 | 0 | 0 | 0 | 0 | 0 | 0 | 0 | 0 | 0 | 14 | 100 | 0 |
| 2014 | 92 | 4 | 4 | 0 | 0 | 0 | 0 | 0 | 0 | 0 | 0 | 26 | 100 | 0 |
| 2015 | 78 | 20 | 2 | 0 | 0 | 0 | 0 | 0 | 0 | 0 | 0 | 46 | 100 | 0 |
| 2016 | 22 | 52 | 22 | 4 | 0 | 0 | 0 | 0 | 0 | 0 | 0 | 23 | 96 | 4 |
| 2017 | 26 | 57 | 13 | 4 | 0 | 0 | 0 | 0 | 0 | 0 | 0 | 23 | 96 | 4 |
| 2018 | 13 | 47 | 33 | 7 | 0 | 0 | 0 | 0 | 0 | 0 | 0 | 15 | 93 | 7 |
| 2019 | 18 | 36 | 36 | 9 | 0 | 0 | 0 | 0 | 0 | 0 | 0 | 11 | 91 | 9 |
| 2020 | 35 | 39 | 23 | 3 | 0 | 0 | 0 | 0 | 0 | 0 | 0 | 66 | 97 | 3 |
| 2021 | 23 | 38 | 20 | 12 | 4 | 2 | 1 | 0 | 0 | 0 | 0 | 204 | 81 | 19 |

Cells in orange: Mode (most frequent value)
In yellow: left and right values to the mode

**Table 6:** Distribution of the percentage of MIC to ISAVUCONAZOLE of *C. parapsilosis* strains received at the SMRL since 2016

| YEAR | MIC (mg/L) |  |  |  |  |  |  |  |  |  |  | N |
| --- | --- | --- | --- | --- | --- | --- | --- | --- | --- | --- | --- | --- |
|  | 0.016 | 0.031 | 0.06 | 0.125 | 0.25 | 0.5 | 1 | 2 | 4 | 8 | >8 |  |
| 2016 | 86 | 14 | 0 | 0 | 0 | 0 | 0 | 0 | 0 | 0 | 0 | 7 |
| 2017 | 75 | 15 | 5 | 0 | 5 | 0 | 0 | 0 | 0 | 0 | 0 | 20 |
| 2018 | 87 | 7 | 7 | 0 | 0 | 0 | 0 | 0 | 0 | 0 | 0 | 15 |
| 2019 | 64 | 18 | 9 | 0 | 0 | 9 | 0 | 0 | 0 | 0 | 0 | 11 |
| 2020 | 30 | 14 | 41 | 9 | 0 | 0 | 0 | 0 | 0 | 0 | 0 | 62 |
| 2021 | 38 | 23 | 29 | 7 | 2 | 1 | 1 | 0 | 0 | 0 | 0 | 196 |

Cells in orange: Mode (most frequent value)
In yellow: left and right values to the mode

The presence of *ERG11* mutations in fluconazole non-susceptible isolates was investigated. The *ERG11* gene of 230 strains from 2020 and 2021, including S (n=34), I (susceptible, increased exposure, n=7) and R (n=189) strains to FLC was sequenced. The *ERG*11 gene was found to be wild-type in all the susceptible strains, and in 4.8% of the FNS isolates Among the latter (n=11), one strain was also susceptible increased exposure (I) and six resistant to voriconazole. The remaining fluconazole-resistant isolates (n=178, 95.2%) harbored the Y132F mutation, which has already been associated to FLC resistance in *C. parapsilosis* (Table 7). In addition, we found that one of the resistant isolates harbored the K143R mutation in *ERG11* gene, which has been detected in azole non-susceptible strains causing monoclonal outbreaks in India [26] and also in combination with the Y132F mutation [11]. This mutation has also been associated with pan-azole resistance in *C. tropicalis* [27]. Another strain harbored the G458S mutation, which has also been related to azole resistance in *Candida parapsilosis* [4, 28]. Finally, many isolates harbored the R398I (data not shown), but this mutation was also found in several susceptible isolates, which suggest that it is not related to FLZ resistance.

**Table 7.** Mutations in the *ERG11* gene found in susceptible, susceptible increased exposure (I) and resistant strains to fluconazole. For each category, we also include the susceptibility profile (S/I/R) for voriconazole. HET: heterozygous; HOMO: homozygous.

| *ERG11*<br>Mutation | FLC<br>Susceptible | FLC<br>Susceptible increased<br>exposure |  |  | FLC Resistant |  |  |
| --- | --- | --- | --- | --- | --- | --- | --- |
|  |  | VOR_S | VOR_I | VOR_R | VOR_S | VOR_I | VOR_R |
| WT | 34 | 1 | 1 | 0 | 5 | 1 | 6 |
| Y132F_HET | 0 | 3 | 1 | 0 | 1 | 17 | 38 |
| Y132F_HOMO | 0 | 0 | 0 | 1 | 5 | 29 | 87 |

Interestingly, in up to 35% of the FNS strains, the Y132F substitution was found in heterozygosis. Thus, we analysed if the triazole MIC distribution differed among homozygous or heterozygous strains. We observed that strains that harbored the Y132F mutation in homozygosis had higher MICs to fluconazole (Geometric Mean = 26.1 mg/L) compared to those strains carrying the mutation in heterozygosis (Geometric Mean = 12.5). A similar situation was found for voriconazole (GM of heterozygous strains = 0.39 mg/L vs GM for homozygous strains = 0.5 mg/L). The Y132F substitution did not have a significant influence on the susceptibility to isavuconazole, posaconazole and itraconazole. Moreover, for these three antifungals, the Y132F mutation in homozygosis tended to result in lower GM than in heterozygous strains (table 8).

**Table 8.**
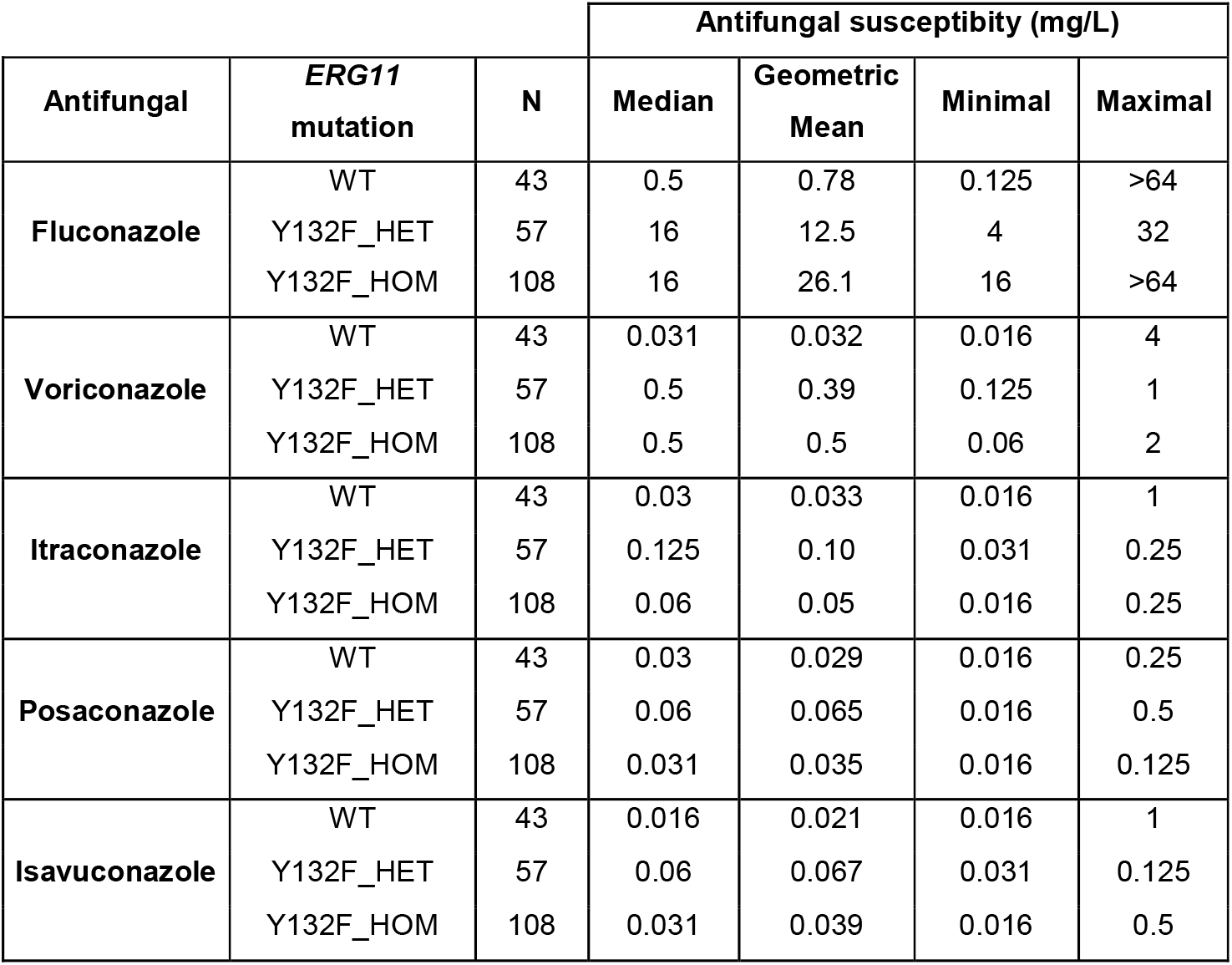
Susceptibility profile of WT or mutant strains harboring the Y132F in homozygosity (HOMO) or heterozygosity (HET).

| Antifungal | *ERG11*<br>mutation | N | Antifungal susceptibility (mg/L) |  |  |  |
| --- | --- | --- | --- | --- | --- | --- |
|  |  |  | Median | Geometric<br>Mean | Minimal | Maximal |
| **Fluconazole** | WT | 43 | 0.5 | 0.78 | 0.125 | >64 |
|  | Y132F_HET | 57 | 16 | 12.5 | 4 | 32 |
|  | Y132F_HOM | 108 | 16 | 26.1 | 16 | >64 |
| **Voriconazole** | WT | 43 | 0.031 | 0.032 | 0.016 | 4 |
|  | Y132F_HET | 57 | 0.5 | 0.39 | 0.125 | 1 |
|  | Y132F_HOM | 108 | 0.5 | 0.5 | 0.06 | 2 |
| **Itraconazole** | WT | 43 | 0.03 | 0.033 | 0.016 | 1 |
|  | Y132F_HET | 57 | 0.125 | 0.10 | 0.031 | 0.25 |
|  | Y132F_HOM | 108 | 0.06 | 0.05 | 0.016 | 0.25 |
| **Posaconazole** | WT | 43 | 0.03 | 0.029 | 0.016 | 0.25 |
|  | Y132F_HET | 57 | 0.06 | 0.065 | 0.016 | 0.5 |
|  | Y132F_HOM | 108 | 0.031 | 0.035 | 0.016 | 0.125 |
| **Isavuconazole** | WT | 43 | 0.016 | 0.021 | 0.016 | 1 |
|  | Y132F_HET | 57 | 0.06 | 0.067 | 0.031 | 0.125 |
|  | Y132F_HOM | 108 | 0.031 | 0.039 | 0.016 | 0.5 |

To investigate if there was any genetic correlation between the FNS strains, we performed a microsatellite-based genotyping of 256 *C. parapsilosis* (from 2019, 2020 and 2021) isolates from 220 different patients and 8 environmental strains, including 81 susceptible, 6 susceptible increased exposure and 168 resistant isolates. Among the susceptible isolates, we included strains from the same hospitals that had resistant strains, but also others not related to these outbreaks. Microsatellite genotyping identified 118 different genotypes. The relationship between the obtained genotypes is illustrated in Fig. 1 and supplemental table 1.

**Figure 1.**
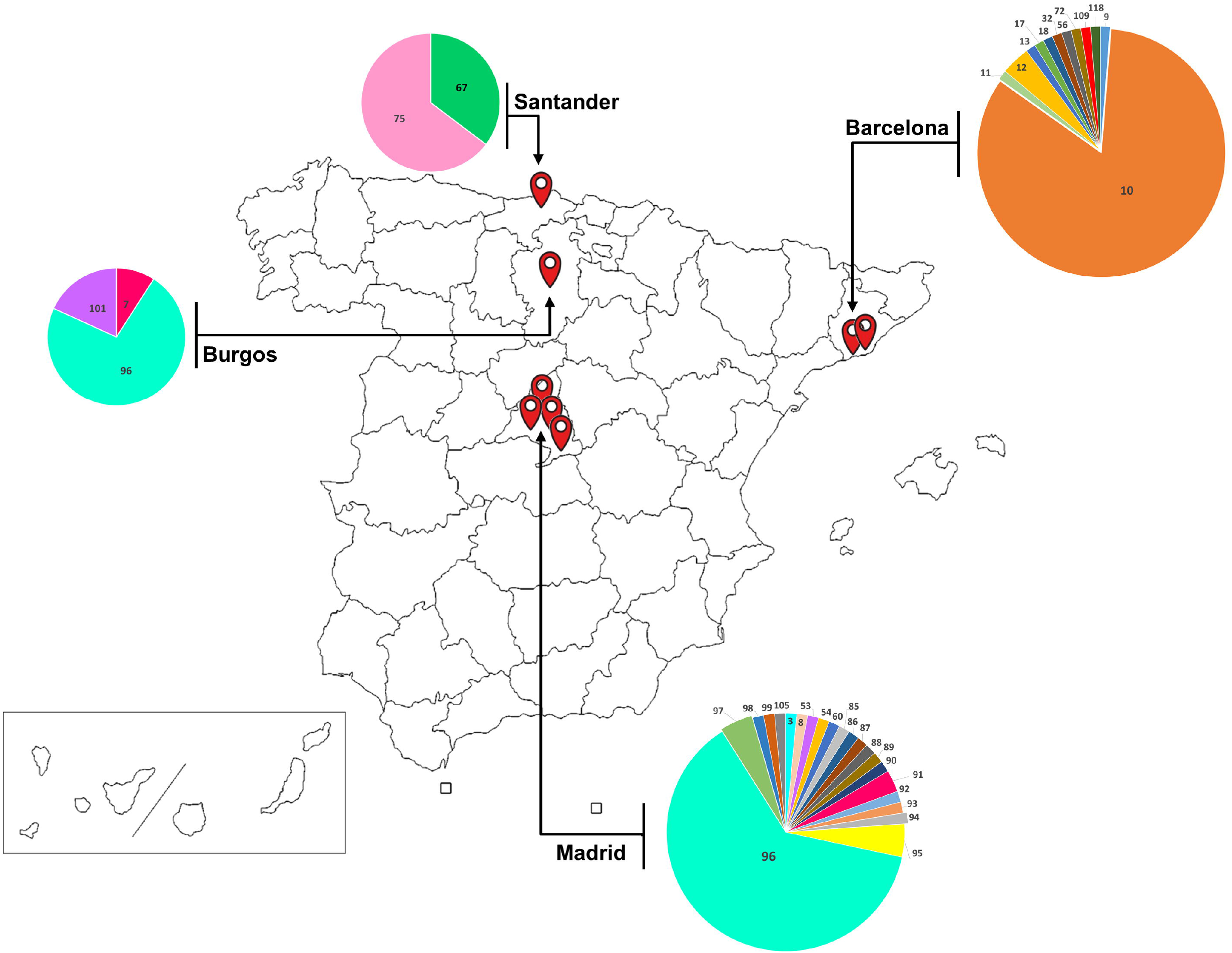
Geographical distribution of the different genotypes of FLZ-resistant isolates. The pie charts denote the distribution of the different genotypes in different tertiary hospitals from different metropolitan areas in Spain.

As compared to the FNS isolates, the genotypic variability was greater among fluconazole-susceptible strains, what could be attributed, in part to the fact that most of the susceptible strains were recovered from unrelated cases.

Remarkably, in the case of contemporary resistant isolates there was a markedly well-defined geographical distribution of genotypes. Genotype 10 was found among strains of two hospitals from the area of Barcelona (Bellvitge and Vall d’Hebron Hospitals), and in an isolate from 2019 stored in the collection and recovered in another center located in the metropolitan area of Barcelona. These two hospitals also shared the closely related genotype 12. Neither genotype 10, nor genotype 12 were found in centers from other regions in Spain. Genotype 96 was found to be highly prevalent among isolates obtained from centers located in Madrid and in Burgos. Genotype 95, despite being much less prevalent, was identified in two centers of the Madrid metropolitan area. Genotypes 67 and 75 were found exclusively in a hospital at the north of Spain (Santander), geographically distant from Madrid and Barcelona. Additionally, fluconazole susceptible strains isolated in the context of another nosocomial outbreak (Universitary Clinic Hospital from Valladolid) were received, displaying genotypes clearly different from the abovementioned and closely related to each other (genotypes 45 to 50). A geographical distribution of the genotypes of the resistant strains is shown in figure 1.

A minimum spanning tree was built, showing that some genotypes have evolved by spontaneous changes in one of the microsatellite markers. The microsatellite analysis showed a distribution of clades that grouped by geographic origin, with resistant strains clustering together (Figure 2).

**Figure 2.**
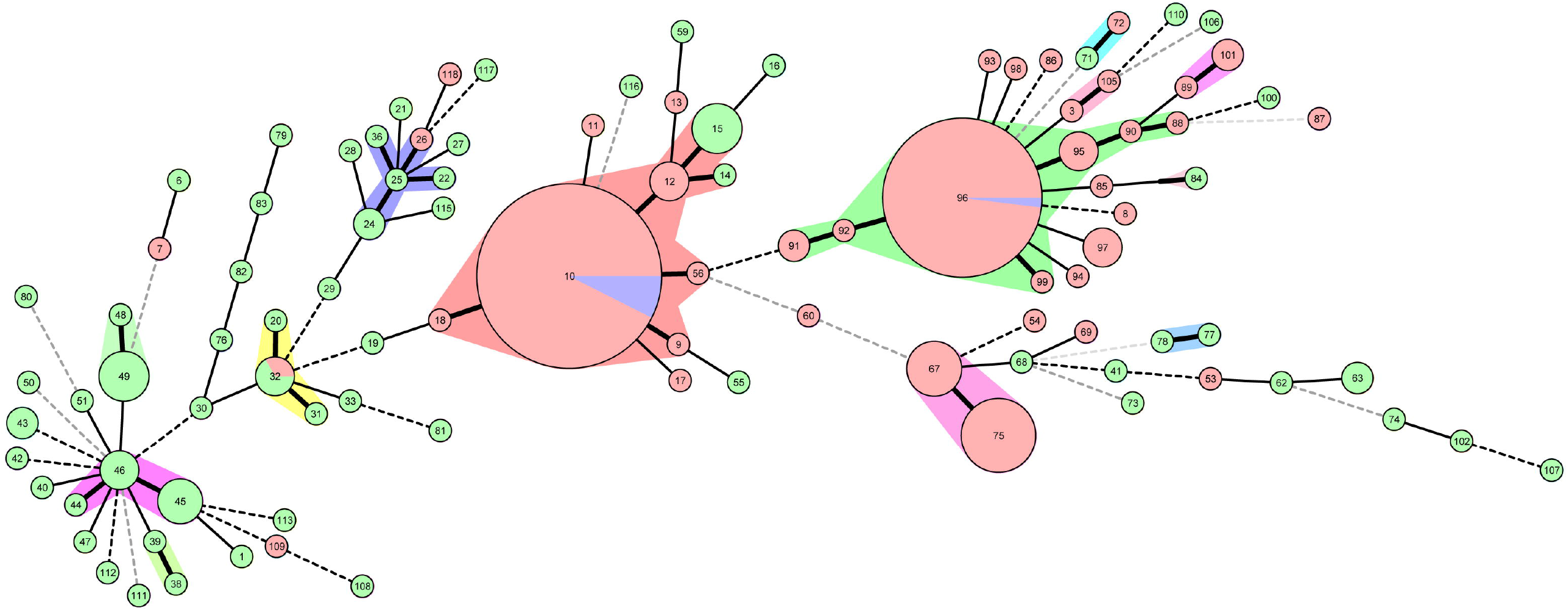
Minimal Spanning Tree showing the genetic proximity of susceptible and FLC-resistant isolates from *Candida parapsilosis*. The numbers denote the genotype identified in each group. Straight bold lines denote groups that only differentiate in one marker. These groups are highlighted with the color shadows in the background. Orange: resistant strains; Green: susceptible strains; Blue: susceptible increased exposure (I) strains. For origin and description of the strains in each genotype, see supplemental table 1.

Our work shows a significant increase in the number of *C. parapsilosis* resistant to fluconazole and voriconazole received at the SMRL from several Spanish hospitals and arising in a relatively short period. This isolates seem to be part of outbreaks that have emerged almost simultaneously in distant cities, and that can be attributed to clones that are shared almost exclusively among geographically close related centers. From these data it cannot be inferred a generalized increment in the fluconazole resistance among Spanish isolates of *C. parapsilosis* since it is not mandatory to inform about all the infections caused by these species. It should be noted that another outbreak of FNS *C. parapsilosis* has been recently described in the Balearic Islands (Son Espases Hospital) [29], which supports the hypothesis that fluconazole resistant strains from *C. parapsilosis* may have emerged and spread in Spain in the last two years. All together, our data is in sharp contrast to what have being described in the several former epidemiological studies that have been carried out in Spain [6, 7, 30, 31], suggesting a new and worrisome change in the epidemiological incidence of FNS *C. parapsilosis* strains.

Recent emergence of FNS isolates in *C. parapsilosis* has been described in other countries in the literature [11-20], so our data supports that the increase of azole resistance in *C. parapsilosis* might be a global problem. In this study, the majority of resistant isolates harbored the Y132F mutation, which has been largely associated in the literature with the appearance of clonal outbreaks. However, we also detected a few isolates that did not have this mutation. For this reason, further studies should be performed to describe all the resistance mechanisms circulating among Spanish hospitals.

At the moment, the reasons for the increase in the incidence of azole-resistant *C. parapsilosis* strains in Spain are unknown, but we hypothesized that this phenomenon may be related to the negative impact that the COVID-19 pandemic has had in Spanish hospitals for several reasons. First, there is a clear temporal correlation between the increase in the number of resistant isolates received at the reference laboratory and clinical impact of the pandemic. Second, the COVID-19 pandemic has resulted in a severe overcrowding of hospitals, and in particular, of Intensive Care Units, along with the necessity of recruiting large numbers of healthcare professionals that were not properly trained in infection control measures. Third, during the pandemic there were changes in personal protective equipment use and the same gloves could have been used between patients [32, 33]. This might have increased the risk of cross-transmission between patients and caused hospital outbreaks. Furthermore, during the pandemic, there has been a significant transfer of patients between different hospitals, which might have contributed to the dispersion of resistant clones between clinical tertiary centres. A similar situation has been described in multicenter studies in India [26], which highlights the ability of FNS isolates to spread and colonize hospital environments. Interestingly, some of the analyzed samples in our work were isolated from environmental origin in the hospital and were also found in clinical samples from the same center. This correlation suggests that *C. parapsilosis* clones might have colonized the hospital surfaces, which increases the risk of recirculating among patients along the time and, in parallel, increases the risk of invasive infections among the most fragile ones. Previous studies, in fact, have shown, not only an increase incidence of *C. parapsilosis* infections in COVID-19 patients [34], but also other fungal diseases, such as Covid Associated Pulmonary Aspergillosis (CAPA) [35-37], mucormycosis [38-40] and *Candida* infections [41, 42] (see reviews in [43, 44]).

The impact of the COVID-19 pandemic and clinical management of the patients does not fully explain why there has been a selection of azole-resistant strains, and why these genetically different resistant strains have emerged almost simultaneously in distant places across Spain. An increase in the use of antimicrobials has been reported since the appearance of the COVID-19 pandemic in some geographical regions [45]. Among azoles, an increase in the use of echinocandins and voriconazole has been reported [45], which might have favoured the selection of fluconazole and voriconazole-resistant *C. parapsilosis*. Another possibility is that resistance to azoles affects virulence traits. In this sense, it has been described that *C. parapsilosis* strains harboring the Y132F mutation in *ERG11* have reduced ability to form biofilms [11], which rises the hypothesis that these strains have a higher ability to spread and disseminate. Furthermore, several studies have associated the incidence of resistant strains with higher mortality of the patients [11, 28], which warrants further studies on the virulence of FLZ non-susceptible *C. parapsilosis* strains. In our case, the clinical management of the patients might have contributed to the selection of pre-existing resistant clones circulating in the hospitals previous to the COVID-19 pandemic [46]. In our case, this idea is supported by the fact that we identified that some of the resistant clones were already present in our collection in samples from 2019. For these reason, it is required to develop future research lines to investigate the genetic proximity of the resistant isolates, and compare them not only between different hospitals, but also to those described in different countries.

Despite the epidemiological limitations and interpretations of our work, we believe that the data herein presented is an indicator of an emerging clinical problem, that is, the selection of azole-resistance in *C. parapsilosis* during the COVID-19 pandemic. We also would like to highlight that the increase in FLZ-resistant isolates in tertiary hospitals in Spain is agreement with the worldwide context, where an increasing number of outbreaks is being reported. We encourage the clinical community to investigate the presence of these clones in the hospital environment, as well as to make an effort to perform susceptibility testing in strains from non-invasive origin (colonization, isolated from hospital surfaces, etc) and to design specific measures to prevent the expansion of the associated resistance mechanisms.

## Data Availability

All data produced in the present study are available upon reasonable request to the authors

## Acknowledgements and Funding

O.Z. was funded by grants SAF2017-86912-R and PID2020-114546RB-I00 from the Spanish Ministry for Science and Innovation. This work was also funded by the National Centre for Microbiology (Instituto de Salud Carlos III) through the Surveillance program of Antifungal Resistance and the Center for Biomedical Research in Network in Infectious Diseases (CIBERINFECT CB21/13/00105 (OZ and LAF) and CB21/13/00009 (M.P-A). L.A-F. was supported by Fondo de Investigación Sanitaria (MPY 117/18 and MPY 305/20).

## Conflict of interest

The authors have no conflict of interest to declare

## Notes

### Competing Interest Statement

The authors have declared no competing interest.

